# Ocular Antibiotic Use in Young Danish Children from 2016 to 2023

**DOI:** 10.1101/2025.02.26.25322849

**Authors:** Heidi Sonne, Anton Pottegård, Katrine Sommerlund, Camilla Flintholm Raft, Helene Kildegaard

## Abstract

**Background:** Ocular antibiotics are frequently prescribed to treat eye infections among young children, despite most cases resolving spontaneously. Daycare policies, parental pressure, and misunderstanding of clinical guidelines may drive overuse. Monitoring prescription trends is essential to guide initiatives to reduce inappropriate use.

**Objectives:** To support an upcoming Danish Choosing Wisely recommendation, we examined trends in ocular antibiotic use among Danish children aged 0–5 years from 2016-2023.

**Methods:** This nationwide descriptive study used individual-level data from the Danish National Prescription Registry. Annual prevalence and incidence rates of antibiotic use were calculated stratified by age, sex, region, and municipality. Treatment episodes per child and prescriber type were also analyzed.

**Results:** During the study period, 568,801 prescriptions for ocular antibiotics were issued to 317,283 children aged 0–5 years. Use declined steadily from 2016-2019. A substantial reduction occurred during the COVID-19 pandemic, followed by a rebound. During 2023, 1-year-olds had the highest rates of use (386 per 1,000 boys; 321 per 1,000 girls), with 29% of children receiving at least one treatment episode. Considerable regional variation in prescribing existed with an incidence rate ratio of 1.45 between the South and Mid Regions of Denmark.

**Conclusion:** Ocular antibiotic use is rising after the COVID-19 pandemic, with the highest use among children aged 1-2 years and with significant regional variation.

## Introduction

Ocular antibiotics are commonly prescribed to treat eye infections. Among young children, around half of acute conjunctivitis cases are bacterial,^1^ but distinguishing between viral and bacterial conjunctivitis can be challenging.^1–3^ Most cases resolve spontaneously without treatment,^1^ yet the prescription of ocular antibiotics may be motivated by parental pressure and daycare or school policies requiring treatment for reentry.^4,5^ Such practices raise concerns about inappropriate use, which can contribute to the development of antibiotic resistance.^6^ To address such issues, Choosing Wisely programs have been launched in over 30 countries, including Denmark, where Choosing Wisely Denmark was established in 2020. The program aims to reduce unnecessary tests, procedures, and treatments in healthcare, including antibiotic overuse.^7,8^ However, making informed recommendations requires detailed knowledge of antibiotic consumption. This study supports an upcoming Danish Choosing Wisely recommendation by analyzing trends in pediatric ocular antibiotic use in Danish children from 2016 to 2023, extending previous work.^9^

## Methods

In this nationwide descriptive drug utilization study, we extracted individual-level data on all redeemed prescriptions for ocular antibiotics issued for children (age <6 years at the time of redemption) from the 1st of January 2016 to the 31st of December 2023.

### Setting

Choosing Wisely Denmark was established as a joint initiative by the umbrella organization of patient associations and scientific societies to identify areas in Danish healthcare where unnecessary tests, procedures, or treatments are performed, some of which may potentially be harmful to patients. The Danish organization has a unique model, where patients and healthcare professionals work together as equals to identify unnecessary practices. In other countries Choosing Wisely is led solely by physicians. Choosing Wisely Denmark develops evidence-based ‘do-not’ recommendations on tests, procedures, or treatments that should be avoided in healthcare.^7^

### Data sources

The Danish National Prescription Registry contains individual-level data on all prescriptions filled by Danish residents at community pharmacies since 1995.^10^ It holds information on the drug dispensing date, the Anatomical Therapeutic Chemical (ATC) classification code, and strength.^10^ All antibiotics, irrespective of the route of administration, require a prescription from a licensed medical provider. We included the following Anatomical Therapeutic Chemical (ATC) groups: S01AA (general antibiotics) and S01AE (fluoroquinolones). Sulfonamides (S01AB) were not marketed in Denmark during the study period and therefore not included.

### Statistical analyses

Four different analyses were performed. First, the annual prevalence proportion of ocular antibiotic use was calculated as the total number of children redeeming at least one prescription for any ocular antibiotic each year divided by the total population of children that year, reported per 1,000 individuals. The prevalence estimates were further stratified by sex, age group at the time of filling the prescription (0–1 and 2–5 years), and antibiotic type, including chloramphenicol (S01AA01), fusidic acid (S01AA13), tobramycin (S01AA12), ciprofloxacin (S01AE03), and other antibiotics. Second, we calculated the annual and monthly incidence rate of antibiotic use as the number of treatment episodes per 1,000 children. Clusters of prescriptions separated by less than 14 days were considered part of the same treatment episode. Incidence were further stratified by sex, age, residential region, and municipality.

Third, the number of antibiotic treatment episodes per child in 2023 was assessed by estimating the proportion of children who received 1, 2, or 3+ treatment episodes using a 365-day look-back period from each child’s birthday in 2023. These results were stratified by 1-year age bins. Finally, we investigated the type of prescriber responsible for each prescription. Analyses were conducted using Stata version 18 (StataCorp, College Station, TX).

## Results

From 2016 to 2023, a total of 571,463 prescriptions for ocular antibiotics were issued to 319,698 children aged 0–5 years. In 2016, the prevalence of ocular antibiotic use was 322 per 1,000 children aged 0-1 years and 128 per 1,000 children aged 2-5 years (**Figure 1A**). Prevalence decreased sharply in 2020, coinciding with the onset of the COVID-19 pandemic, before gradually increasing in subsequent years. By 2023, the prevalence reached 236 per 1,000 children aged 0-1 years and 143 per 1,000 children aged 2-5 years.

**Figure 1.**
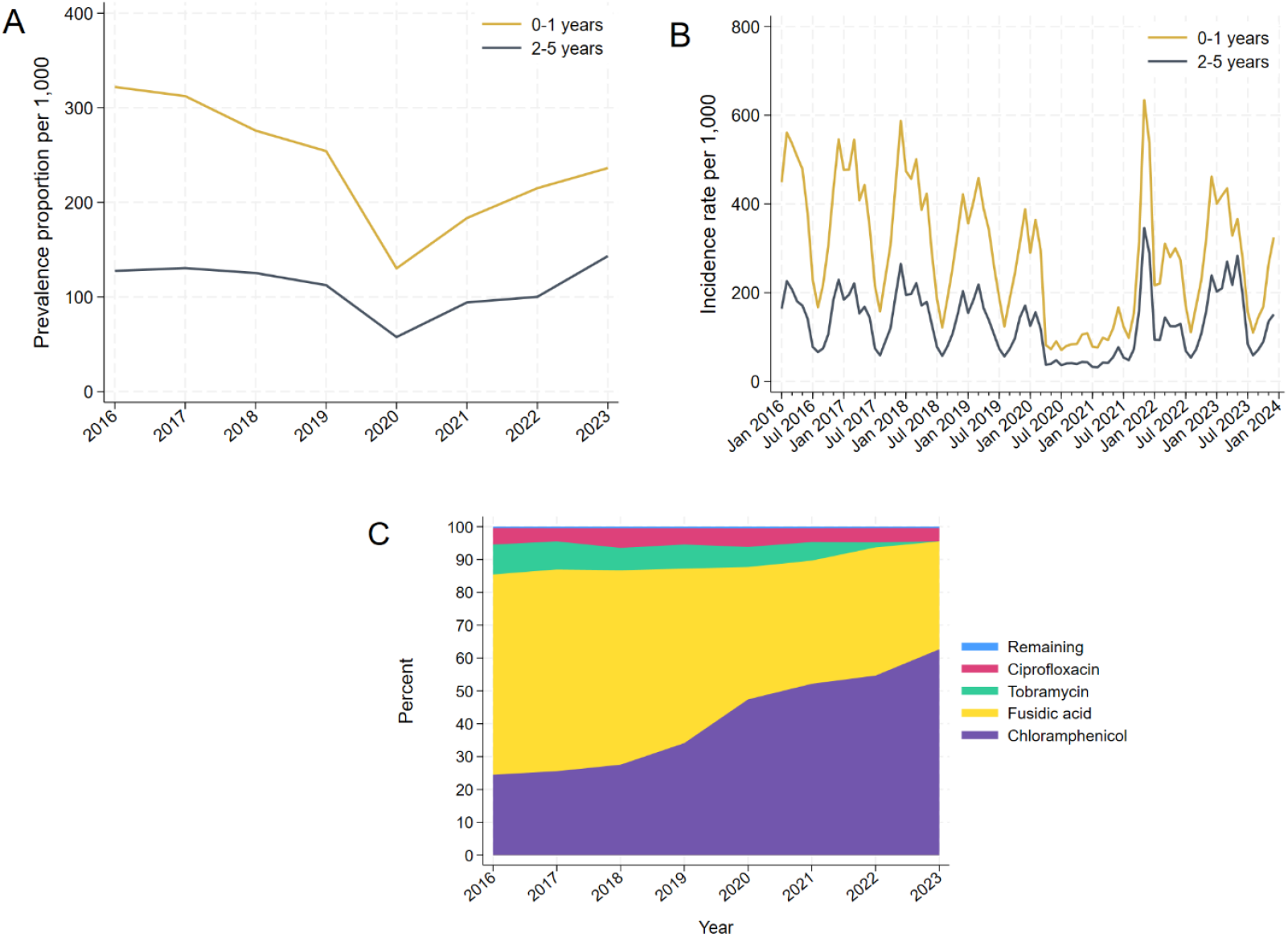
Prevalence and incidence rates of ocular antibiotic use among Danish children from 2016 to 2023. **A**. Annual prevalence of children aged 0–1 years and 2–5 years redeeming at least one prescription for ocular antibiotics per 1,000 children. **B**. Monthly incidence rates of ocular antibiotic treatment episodes per 1,000 children. **C**. Proportional use of the most commonly prescribed antibiotics from 2016 to 2023.

A similar trend was observed in the incidence, with a marked decline during the pandemic, with a plateau between April 2020 and September 2021 (**Figure 1B**). Following this, a large peak occurred, particularly among children aged 0-1 years. Throughout the study period, incidence rates were consistently higher for children aged 0-1 years than those for children aged 2-5 years.

Seasonal variations were evident, with peaks in the winter months and declines in summer. Of all treatment episodes, 86% were prescribed by a general practitioner.

The distribution of antibiotic types shifted significantly over the study period (**Figure 1C**). Fusidic acid was the most commonly prescribed antibiotic in 2016, accounting for approximately 64% of all prescriptions, but the proportion decreased to 33% by 2023. In contrast, chloramphenicol became the dominant antibiotic by 2021, rising from 22% of prescriptions in 2016 to 62% in 2023. The use of tobramycin declined steadily, representing 8.7% of prescriptions in 2016 but not used by 2023. The use of ciprofloxacin remained stable at around 9% throughout the study period.

The incidence rates of ocular antibiotic use per 1,000 children varied by age and sex (**Figure 2A**). In 2023, boys had higher incidence rates compared to girls across all age groups, although differences were attenuated with increasing child age. The highest incidence rate was observed in children aged 12-15 months with an IR of 460-500 per 1,000 for boys and 370-430 per 1,000 for girls. The incidence rate decreased with age, reaching 109 per 1,000 for boys and 105 per 1,000 for girls by age 5. The number of antibiotic treatment episodes per child in 2023 also varied by age (**Figure 2B**). In the second year of life, 29% of children received at least one treatment episode, with 22% receiving only one, 5.2% receiving two, and 1.4% having three or more treatment episodes. The fewest treatment episodes were observed for children aged 5 years old.

**Figure 2.**
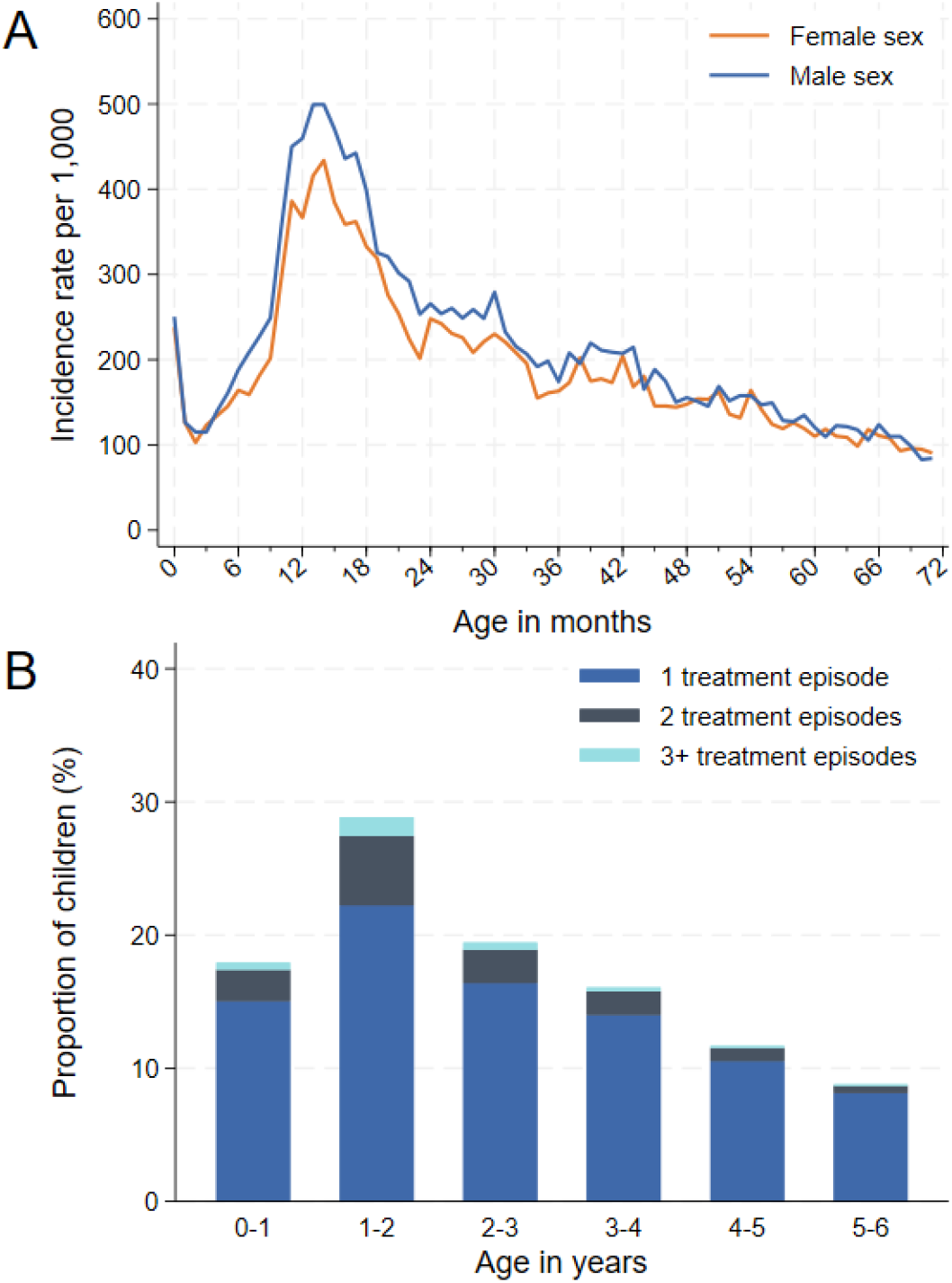
Stratified incidence rates and treatment episodes of ocular antibiotics in 2023. **A**. Incidence rates of ocular antibiotic treatment episodes per 1,000 children by sex and age in 1- month bins. **B**. Proportion of children receiving 1, 2, or 3+ antibiotic treatment episodes in 2023, stratified by 1- year age bins.

Prescribing of ocular antibiotics varied considerably across Denmark. In 2023, the South and Zealand Regions had the highest incidence rates (252 and 223 per 1,000 children, respectively), while the Mid Region had the lowest (174 per 1,000), corresponding to an incidence rate ratio of 1.45. This variation appeared to be driven primarily by differences at the municipal level (**Figure 3**). The highest incidence rates were concentrated in the southern part of Jutland and select areas of Zealand, with rates reaching approximately 300 per 1,000 children. In contrast, municipalities in northern and central Denmark had significantly lower rates, with some below 150 per 1,000 children.

**Figure 3.**
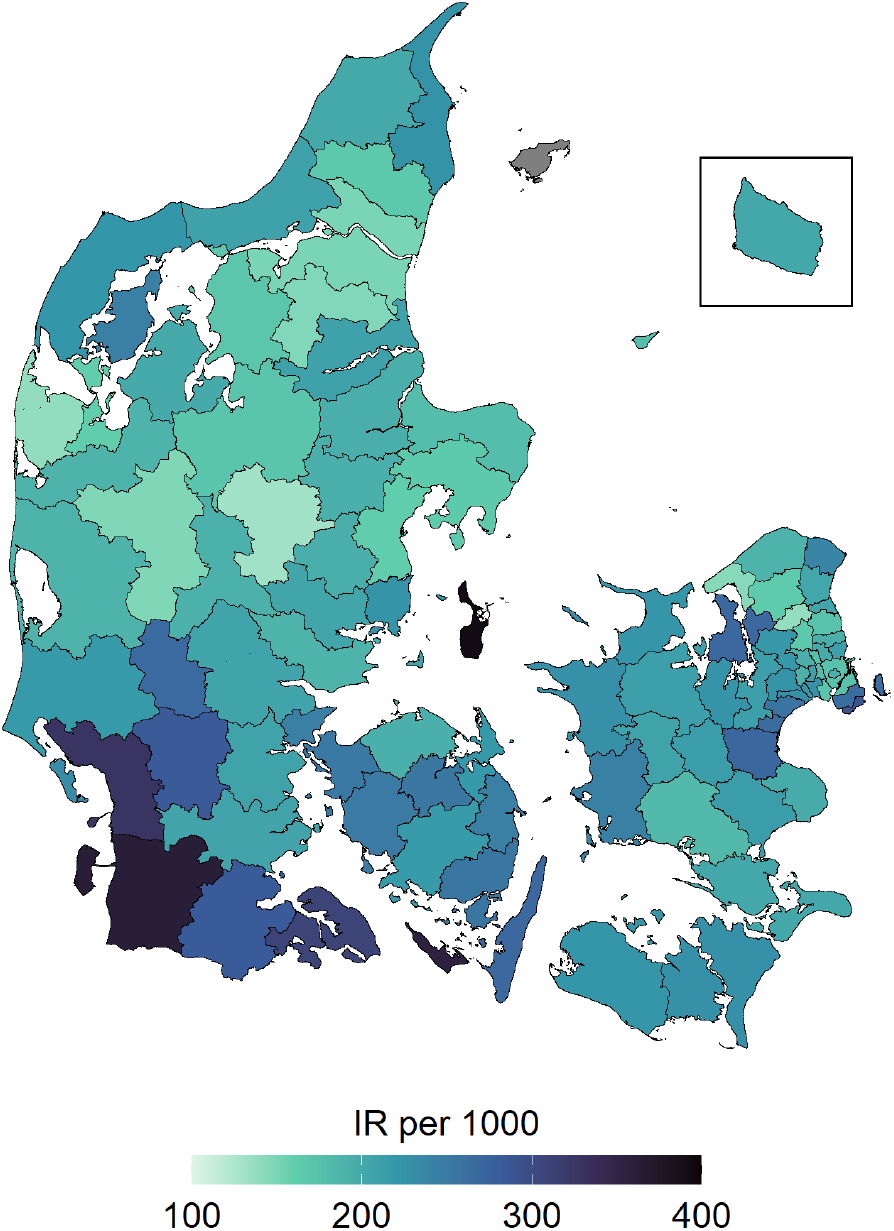
Municipal incidence rates of ocular antibiotics in 2023. Incidence rates of ocular antibiotic treatment episodes per 1,000 children by Danish municipalities in 2023.

## Discussion

This study provides contemporary data on trends in the use of ocular antibiotics among Danish children aged 0-5 years. We observed a significant decline in both prevalence and incidence rates of ocular antibiotic use during the COVID-19 pandemic, followed by a resurgence in the post-pandemic period. Chloramphenicol has replaced fusidic acid as the most prescribed antibiotic, while the use of tobramycin has disappeared. Antibiotic use was highest among children aged 0-1 years and varied considerably across regions, with the South and Zealand Regions showing the highest incidence rates.

The strengths of this study include the use of nationwide registry data, which ensures comprehensive coverage and eliminates selection bias. Additionally, the data capture all prescriptions filled at community pharmacies, regardless of whether they were issued by hospitals, general practice, or out-of-hours primary care.^10^ However, limitations include the inability to assess whether the ocular antibiotics were used as prescribed and the absence of clinical information which enables evaluation of prescribing appropriateness.

Our results show a marked decrease in the use of ocular antibiotics during 2020 and 2021, coinciding with COVID-19 restrictions. After restrictions were lifted, we observed a sharp increase in prevalence and incidence, likely reflecting a rebound in eye infections as exposure to pathogens resumed.^11,12^ A similar trend was observed in England, reporting a decrease in antibiotic prescriptions during the pandemic, followed by a resurgence.^13^ Despite this rebound, prevalence and incidence rates in 2023 remained below 2016 levels which align with prior findings from 2000- 2015 showing a general decline in ocular antibiotic prescriptions.^9^

Compared to other Scandinavian countries, Denmark continues to have a higher prevalence of ocular antibiotic use.^9^ In Norway, the prevalence of ocular antibiotics among children aged 0–4 years was lower but comparable to Denmark (153 per 1,000 children), with a steady decline observed from 2016 to 2023.^14^ In contrast, Sweden had markedly lower prevalence levels in 2023 (40 per 1,000 children),^15^ which accounts for only about a quarter of those in Denmark, suggesting substantial potential for reducing antibiotic use in Denmark.

Within Denmark, we observed significant regional and municipal variations in prescribing patterns. These differences may reflect variations in clinical practice, access to healthcare services, or regional guidelines. Regional efforts to promote appropriate antibiotic use, such as educational campaigns, have shown success in reducing prescriptions and could be expanded to address these disparities.^16^ Initiatives such as Choosing Wisely Denmark have the potential to further reduce ocular antibiotic overuse by developing a specific ‘do-not’ recommendation on this issue and increasing awareness not only among clinicians but also within the general population.

We consistently observed higher incidence rates among children aged 0-1 years compared to children aged 2-5 years, aligning with the age at which Danish children typically begin daycare, likely increasing their exposure to pathogens. Daycare policies that require treatment for eye infections before reentry could further drive antibiotic prescriptions in this age group,^4^ despite national guidelines recommending antibiotics only for severe symptoms of bacterial conjunctivitis.^17^

Parental expectations and limited knowledge about antibiotics likely also contribute to overuse.^5^ In the Capital Region, 74% of general practitioners reported direct parental requests for antibiotics, and 88% experienced indirect pressure.^16,18^ Addressing these factors through education and awareness initiatives could help reduce unnecessary antibiotic use.

## Conclusion

This study highlights important trends in ocular antibiotic use among young children in Denmark from 2016 to 2023 with the aim of supporting a Choosing Wisely Denmark recommendation on the same topic. A steady decline in use was observed from 2016 to 2019, followed by a temporary reduction during the COVID-19 pandemic and a subsequent rebound. These findings underscore the need for continued efforts to promote appropriate use. Addressing parental expectations, standardizing national guidelines, and reducing regional disparities could help optimize prescribing practices and limit unnecessary antibiotic use, thereby contributing to antimicrobial stewardship.

## Ethics

According to Danish law, studies based solely on register data do not require ethical approval.

## Data availability statement

Individual-level data cannot be shared by the authors owing to Danish data protection regulations. De-identified data can be made available for authorized researchers after application to Forskerservice at the Danish Health Data Authority.

